# Renin-angiotensin system inhibitors and susceptibility to COVID-19 in patients with hypertension: a propensity score-matched cohort study in primary care

**DOI:** 10.1101/2020.09.17.20196469

**Authors:** Shamil Haroon, Anuradhaa Subramanian, Jennifer Cooper, Astha Anand, Krishna Gokhale, Nathan Byne, Samir Dhalla, Dionisio Acosta-Mena, Thomas Taverner, Kelvin Okoth, Jingya Wang, Joht Singh Chandan, Christopher Sainsbury, Dawit Tefra Zemedikun, G. Neil Thomas, Dhruv Parekh, Tom Marshall, Elizabeth Sapey, Nicola J Adderley, Krishnarajah Nirantharakumar

**Affiliations:** Institute of Applied Health Research, University of Birmingham, Birmingham, UK; Cegedim Health Data, Cegedim Rx, London, UK; The Health Improvement Network (THIN), London, UK; Department of Diabetes, Gartnavel General Hospital, NHS Greater Glasgow and Clyde, UK; Birmingham Acute Care Research Group, Institute of Inflammation and Ageing, University of Birmingham, Birmingham, UK; Department of Acute Medicine, Queen Elizabeth Hospital Birmingham, UK; PIONEER, The Health Data Research UK Hub in acute care; Department of Diabetes and Endocrinology, University Hospitals Birmingham NHS Foundation Trust, Birmingham, UK; Midlands Health Data Research UK, Birmingham, UK

## Abstract

**Introduction:** A significant proportion of patients with Coronavirus Disease-19 (COVID-19) have hypertension and are treated with renin-angiotensin system (RAS) inhibitors, namely angiotensin-converting enzyme I inhibitors (ACE inhibitors) or angiotensin II type-1 receptor blockers (ARBs). These medications have been postulated to influence susceptibility to Severe Acute Respiratory Syndrome Coronavirus-2 (SARS-CoV-2). The objective of this study was to assess a possible association between prescription of RAS inhibitors and the incidence of COVID-19 and all-cause mortality.

**Methods:** We conducted a propensity-score matched cohort study to assess the incidence of COVID-19 among patients with hypertension who were prescribed ACE inhibitors or ARBs compared to patients treated with calcium channel blockers (CCBs) in a large UK-based primary care database (The Health Improvement Network). We estimated crude incidence rates for confirmed/suspected COVID-19 among those prescribed ACE inhibitors, ARBs and CCBs. We used a Cox proportional hazards model to produce adjusted hazard ratios for COVID-19 comparing patients prescribed ACE inhibitors or ARBs to those prescribed CCBs. We further assessed all-cause mortality as a secondary outcome and a composite of accidents, trauma or fractures as a negative control outcome to assess for residual confounding.

**Results:** In the propensity score matched analysis, 83 of 18,895 users (0.44%) of ACE inhibitors developed COVID-19 over 8,923 person-years, an incidence rate of 9.3 per 1000 person-years. 85 of 18,895 (0.45%) users of CCBs developed COVID-19 over 8,932 person-years, an incidence rate of 9.5 per 1000 person-years. The adjusted hazard ratio for suspected/confirmed COVID-19 for users of ACE inhibitors compared to CCBs was 0.92 (95% CI 0.68 to 1.26). 79 out of 10,623 users (0.74%) of ARBs developed COVID-19 over 5010 person-years, an incidence rate of 15.8 per 1000 person-years, compared to 11.6 per 1000 person-years among users of CCBs. The adjusted hazard ratio for suspected/confirmed COVID-19 for users of ARBs compared to CCBs was 1.38 (95% CI 0.98 to 1.95). There were no significant associations between use of ACE inhibitors or ARBs and all-cause mortality, compared to use of CCBs. We found no evidence of significant residual confounding with the negative control analysis.

**Conclusion:** Current use of ACE inhibitors was not associated with the risk of suspected or confirmed COVID-19 whereas use of ARBs was associated with a statistically non-significant 38% relative increase in risk compared to use of CCBs. However, no significant associations were observed between prescription of either ACE inhibitors or ARBs and all-cause mortality during the peak of the pandemic.

## Introduction

A novel strain of coronavirus, Severe Acute Respiratory Syndrome Coronavirus-2 (SARS-CoV-2) was first detected in December 2019 in the district of Wuhan, China. This infection was found to cause a severe respiratory illness, termed COVID-19, which was associated with the development of acute respiratory distress syndrome (ARDS), particularly in older male adults, in those with obesity and comorbidities, and those from Black and Minority Ethnic backgrounds and low socioeconomic status.(1) The virus has caused a global pandemic that has crippled health systems and economies. As of 24th August 2020, SARS-CoV-2 was estimated to have infected over 23 million people and caused over 800,000 deaths.(2)

Early on in the pandemic, a number of case series of patients with COVID-19 in China indicated a high prevalence of hypertension among those affected.(3)(4) Patients with hypertension appeared to have a threefold increase in the odds of mortality from COVID-19 compared to those without.(5) It is unclear whether this association was causal, and if so whether hypertension or antihypertensive drugs increased the risk of adverse outcomes from COVID-19. The renin angiotensin system (RAS) inhibitors, angiotensin-converting enzyme I inhibitors (ACEIs) and angiotensin II type-1 receptor blockers (ARBs), were specifically postulated to be involved in the pathogenesis of SARS-CoV-2.(6)

SARS-CoV-2 enters human cells using the angiotensin-converting enzyme 2 (ACE-2) receptor, which is expressed in epithelial cells in human organs, including type II alveolar cells in the lungs as well as the cardiovascular system, kidneys, adrenal glands, brain, uterus and skin.(7)(8)(9) Experimental studies have suggested that use of ACE inhibitors and ARBs can upregulate ACE-2 receptor expression in the cardiovascular and renal system.(10) RAS inhibitors are frequently used by patients with hypertension, diabetes, chronic kidney disease, and cardiovascular disease, all groups that are at increased risk from COVID-19.(1) However, it remains unclear whether this association is due to the underlying cardiometabolic state associated with these conditions or the pharmacological agents used to treat them. Furthermore, the pathways within the renin-angiotensin system are complex and ACE inhibitors and ARBs may theoretically be protective because they increase concentrations of ACE-2 and angiotensin (1-7), which have been shown to be protective in lung injury models.(11)

The relationship between ACE inhibitors and ARBs and risk of COVID-19 need to be clarified as a large proportion of patients with hypertension, type 2 diabetes, heart failure, and chronic kidney disease, all of which are considered risk factors for COVID-19, are currently prescribed these drugs. In the absence of this evidence, it would not be appropriate to withdraw or switch these drugs as they are known to be cardioprotective and renoprotective. The Council on Hypertension of the European Society of Cardiology highlighted the lack of evidence supporting harmful effects of ACE inhibitors and ARBs in the context of COVID-19 early on in the pandemic.(12) Pharmacoepidemiological studies are needed to address this evidence gap.(13)

One of the challenges with understanding the relationship between COVID-19 susceptibility and use of RAS inhibitors is the difficulty in disentangling the independent effects of comorbidities, such as cardiovascular disease, from exposure to drugs that are used to treat those comorbidities. This can potentially introduce a number of biases. For example, patients may be prescribed a RAS inhibitor over an alternative antihypertensive drug such as a calcium channel blocker because of their age or ethnic group, both of which are risk factors for severe COVID-19. Similarly, individuals may be prescribed RAS inhibitors for a range of indications including hypertension, chronic kidney disease, diabetes with albuminuria, ischaemic heart disease and heart failure, all of which are also associated with the severity of COVID-19. Not accounting for these differences potentially introduces confounding by indication bias, resulting in associations between COVID-19 and exposure to drugs of interest being incorrectly attributed to drug exposures rather than the presence of underlying risk factors.(14)

Several epidemiological studies have attempted to investigate the association between RAS inhibitors and COVID-19 susceptibility. One of the earliest published studies was a case control study conducted in Lombardy, Italy.(15) This looked at 6272 patients with confirmed SARS-CoV-2 infection and compared them to 30,759 controls matched on age, sex, and residential municipality. The study found no association between prescription of RAS inhibitors and COVID-19 on adjusted analyses. However, participants with a range of health conditions relevant to the prescription of RAS inhibitors were included in this analysis, which leaves open the possibility of confounding by indication bias. In a later retrospective cohort study and nested case control study of patients with hypertension in Denmark, investigators also found no significant association between use of RAS inhibitors and COVID-19 susceptibility, severity or mortality.(16) However, this study also included patients with a range of conditions that could also be indications for RAS inhibitors, again introducing the potential for confounding by indication bias.

The ideal study design to answer this question would be a randomised controlled trial comparing RAS inhibitors to comparator drugs. However, such a trial would likely be unfeasible given the widespread use of these drugs in everyday clinical practice. However, one approach to disentangling the independent relationship between COVID-19 susceptibility and exposure to RAS-inhibitors is to study patients with hypertension while excluding those with other indicator conditions such as cardiovascular or chronic kidney disease, and comparing the incidence of COVID-19 among similar patients who have received a RAS inhibitor to those who have received an active comparator drug such as a calcium channel blocker (CCB). This approach would more adequately control for differences in risk by underlying condition and indication for RAS inhibitor prescription, and thus reduce potential confounding by indication bias. The objective of this study is to assess whether there is an independent association between the use of RAS-inhibitors and incidence of COVID-19 and all-cause mortality among patients with hypertension.

## Methods

### Study design

This is a propensity score-matched cohort study with active comparators, using routine primary care data.

### Data source

We used data from The Health Improvement Network database. This is a large database of primary care records from UK general practices that use Vision electronic health record software. It includes data for approximately 14 million patients (2 million active patients) at over 640 primary care practices. It includes coded data on patient demographics, diagnoses, primary care prescriptions, consultations and investigations. Practices contributing data to primary care as of 30^th^ Jan 2020 (index date) were eligible for inclusion if they had shown acceptable mortality reporting and had the Vision system installed on or before 30^th^ Jan 2019. Investigators had direct access to an up-to-date extract of the data source.

### Study population

Adults aged 18 years and older with a diagnosis of hypertension and who were registered with an eligible general practice before 30^th^ Jan 2019 were included. In our primary analysis, we excluded patients with heart failure, diabetes, cardiovascular disease (ischaemic heart disease, transient ischaemic attack, stroke, and peripheral vascular disease), and chronic kidney disease (including patients with an estimated glomerular filtration rate <30/min/1.73m^2^) as these comorbidities represented alternative indications for RAS-inhibitors. We also excluded all patients who were pregnant during the index date or had contraindications to the exposure drugs (e.g. hypersensitivity to ACE inhibitors).

### Exposed and comparator groups

We derived three main cohorts of patients defined by their prescription of one of three antihypertensive drugs - angiotensin-converting enzyme inhibitors (ACE inhibitors) and angiotensin II receptor blockers (ARBs), which were the two exposure drugs of interest, and calcium channel blockers (CCBs), which was the active comparator. Drug prescriptions were ascertained using recorded British National Formulary (BNF) codes.

In the primary analysis, all three cohorts were mutually exclusive and patients having a concurrent prescription of any two of the three medications were excluded. To avoid any residual effect of any of the other two medications in their respective cohorts, those with a preceding prescription of any of the other two medications after 30^th^ October 2019 (3 month washout period) were excluded. However, patients were still included if they had a concurrent prescription of other antihypertensive classes (e.g. diuretics and beta-blockers), but these variables were used to propensity score match the exposure and comparator cohort and further adjusted for in the outcome analysis.

### Matching

We estimated propensity scores for prescription of the treatment of interest (ACE inhibitor/ARB) using logistic regression, including the covariates listed below. Matched paired exposure groups (ACE inhibitors vs CCBs and ARBs vs CCBs) were created after performing 1:1 propensity score matching using the nearest-neighbour algorithm, considering callipers of width equal to 0.2. We matched without replacement. We assessed the covariate balance of the matched groups by calculating the standardized absolute mean difference (SMD), considering SMD below 0.10 a balanced covariate.

### Outcomes

The primary outcome was a composite of confirmed or suspected diagnosis of COVID-19 recorded using the clinical (Read) codes listed in the supplementary table. We also assessed all-cause mortality as a secondary outcome. A negative control was used to assess for residual confounding.(17) This was a composite of accidents, trauma or fractures, which was chosen on the basis that we did not expect it to be differentially associated with either the drugs of interest or the outcome.

### Follow-up period

Patients were followed up from the 30^th^ January 2020 until the earliest of the following: recording of the outcome (as defined above), death, patient left practice/dataset, practice ceased contributing to the database, and study end (22nd July 2020). The latest available baseline covariate data recorded before 30^th^ January 2020 were obtained. We retrospectively captured outcome records available until the study end date.

### Covariates

Baseline covariates were extracted for propensity score matching and model adjustment, which included:

1. Sociodemographic characteristics - age and sex.
2. Lifestyle risk factors and metabolic profile: smoking status, alcohol consumption, body mass index (BMI), blood pressure, total cholesterol, high density lipoprotein (HDL), and estimated glomerular filtration rate (eGFR).
3. Duration of hypertension and age of hypertension diagnosis.
4. Presence of comorbid conditions including those listed as high risk for COVID-19 (12, 13): chronic respiratory disease (including severe asthma and COPD), atrial fibrillation, rheumatoid arthritis, cancers (excluding skin cancer), haematological conditions (including haematological malignancies) and immunosuppressive conditions (including immunodeficiency, use of immunosuppressive drugs, chemotherapy and radiotherapy, antibody treatment for cancer, and solid organ transplant).
5. Concurrent prescriptions for thiazide diuretics, potassium diuretics, alpha-adrenoceptor blockers, beta-adrenoceptor blockers, other antihypertensives, statins, and anticoagulants, as defined based on BNF chapters.

### Sample Size

The study sample size was not determined by an a priori sample size calculation. Rather, we included all patients meeting the study eligibility criteria. There have already been over 300,000 confirmed COVID-19 diagnoses recorded in the UK general population. Considering the prevalence of hypertension to be 30%,(18) and the THIN database constituting more than 2 million active patients, we expected to have sufficient power to detect differences in the incidence rates of the primary outcome. We included all current users of the exposure drugs as described in the section above, minimizing selection bias by using the maximum sample size available.

### Statistical Analysis

We used basic descriptive statistics to summarize the characteristics of the patients in each of the prescription cohorts before and after propensity score matching. Crude incidence rates of each outcome were calculated with 95% confidence intervals (CIs). In the primary analysis, we applied a Cox proportional hazards regression model to determine crude and adjusted hazard ratios (HR) comparing pairs of treatment groups in patients with hypertension. The models were adjusted for the covariates listed above. The Cox proportional hazards assumption was tested using Schoenfeld’s residuals test and log-log plots.

### Sensitivity analyses

We repeated the primary analyses comparing outcomes for patients who used ACE inhibitors with or without CCBs to those prescribed CCBs without a RAS inhibitor. We did the same with patients prescribed ARBs with or without a CCB to those prescribed a CCB. This enabled the sample size to be increased while still assessing the additional effect of the exposure drug over and above the active comparator.

The primary analysis was also repeated including patients with diabetes mellitus, ischemic heart disease, stroke or TIA and peripheral vascular disease, mirroring the inclusion criteria used by Fosbøl et al.(16) We also repeated the analysis further including patients with hypertension, CKD and heart failure, as was done in the study by Mancia et al(19)(19).(15) This allowed us to assess the potential effect of confounding by indication bias introduced by including patients with different indications for the exposure drugs.

### Missing data

Continuous variables such as age, BMI, and total cholesterol were grouped into clinically meaningfully categories. Missing values for smoking status and other categorical variables were treated as a separate missing categorical variable. The absence of a record of any diagnosis (e.g. hypertension, renal disease) was taken to indicate the absence of these conditions.

### Ethical approval

The THIN data collection scheme and research carried out using THIN data were approved by the NHS South-East Multicentre Research Ethic Committee in 2003. Under the terms of the approval, studies must undergo independent scientific review. Approval for this study was obtained from the THIN Scientific Review Committee in June 2020 (SRC protocol reference 20-003-R2).

## Results

### Population selection

Before matching, there were 31,194 individuals with a prescription for ACE inhibitors, 13,377 with a prescription for ARBs, and 27,500 with a prescription for CCBs at the index date. After matching, there were 18,895 patients in each arm of the ACE inhibitor and CCB paired cohorts and 10,623 in each arm of the ARB and CCB paired cohorts (Figure 1).

**Figure 1:**
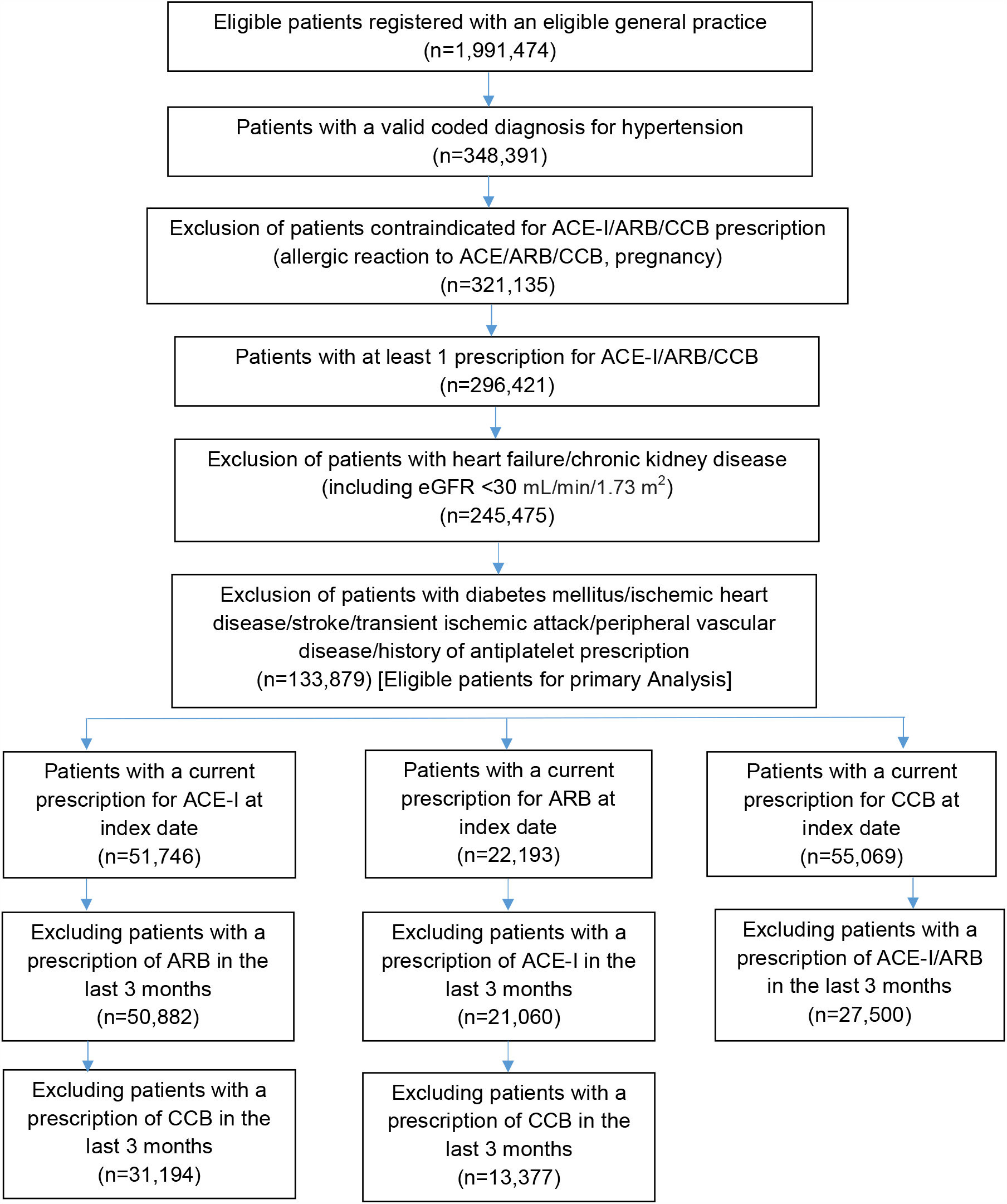
Number of subjects at each stage of the study.

### Study participants

#### ACE inhibitors versus calcium channel blockers

Before matching, the mean age of users of ACE inhibitors and CCBs was 60.8 years and 67.4, respectively (Table 1). The proportion that were male was slightly higher for users of ACE inhibitors than users of CCBs (48.8% vs 45.6%, respectively). A similar proportion were current smokers but a greater proportion of users of ACE inhibitors were overweight or obese compared to users of CCBs (77.7% vs 69.1%, respectively).

**Table 1:** Baseline characteristics of the primary analysis cohorts

|  | Unmatched |  | Propensity score matched |  | Unmatched |  | Propensity score matched |  |
| --- | --- | --- | --- | --- | --- | --- | --- | --- |
| Primary Analysis Cohort | ACE-I<br>(n=31,194) | CCB<br>(n=27,500) | ACE-I<br>(n=18,895) | CCB<br>(n=18,895) | ARB<br>(n=13,377) | CCB<br>(n=27,500) | ARB<br>(n=10,623) | CCB<br>(n=10,623) |
| Age, mean (SD) | 60.8 (11.7) | 67.4 (10.8) | 64.7 (11.2) | 63.4 (9.6) | 63.8 (11.3) | 67.4 (10.8) | 65.0 (11.1) | 64.55 (11.5) |
| Age categories, n (%) |  |  |  |  |  |  |  |  |
| 18 - 30 years | 123 (0.4) | 28 (0.1) | 34 (0.2) | 28 (0.2) | 23 (0.2) | 28 (0.1) | 14 (0.1) | 26 (0.2) |
| 30 - 40 years | 875 (2.8) | 309 (1.1) | 213 (1.1) | 308 (1.6) | 206 (1.5) | 309 (1.1) | 118 (1.1) | 238 (2.2) |
| 40-50 years | 4466 (14.3) | 1302 (4.7) | 1445 (7.7) | 1297 (6.9) | 1298 (9.7) | 1302 (4.7) | 834 (7.9) | 865 (8.1) |
| 50-60 years | 10214 (32.7) | 4836 (17.6) | 5018 (26.6) | 4694 (24.8) | 3556 (26.6) | 4836 (17.6) | 2623 (24.7) | 2472 (23.3) |
| 60-70 years | 8448 (27.1) | 9521 (34.6) | 5879 (31.1) | 7969 (42.2) | 4157 (31.1) | 9521 (34.6) | 3388 (31.9) | 3497 (32.9) |
| 70-80 years | 5380 (17.3) | 8376 (30.5) | 4666 (24.7) | 3974 (21.0) | 3097 (23.2) | 8376 (30.5) | 2703 (25.4) | 2617 (24.6) |
| >80 years | 1688 (5.4) | 3128 (11.4) | 1640 (8.7) | 625 (3.3) | 1040 (7.8) | 3128 (11.4) | 943 (8.9) | 908 (8.6) |
| Sex (Male), n (%) | 15213 (48.8) | 12,547 (45.6) | 8870 (46.9) | 9243 (48.9) | 5379 (40.2) | 12547 (45.6) | 4628 (43.6) | 4463 (42.0) |
| Smoker categories, n (%) |  |  |  |  |  |  |  |  |
| Non Smoker | 17968 (57.6) | 15514 (56.4) | 10725 (56.8) | 10668 (56.5) | 8390 (62.7) | 15514 (56.4) | 6384 (60.1) | 6473 (60.9) |
| Ex-Smoker | 9073 (29.1) | 8329 (30.3) | 5677 (30.0) | 5700 (30.2) | 3865 (28.9) | 8329 (30.3) | 3186 (30.0) | 3137 (29.5) |
| Smoker | 3962 (12.7) | 3483 (12.7) | 2402 (12.7) | 2431 (12.9) | 1058 (7.9) | 3483 (12.7) | 1005 (9.5) | 967 (9.1) |
| Missing | 191 (0.6) | 174 (0.6) | 91 (0.5) | 96 (0.5) | 64 (0.5) | 174 (0.6) | 48 (0.5) | 46 (0.4) |
| Drinker Categories, n (%) |  |  |  |  |  |  |  |  |
| Non-drinker | 4716 (15.1) | 4186 (15.2) | 2874 (15.2) | 2856 (15.1) | 2013 (15.1) | 4186 (15.2) | 1587 (14.9) | 1508 (14.2) |
| Drinker without excess | 11716 (37.6) | 10281 (37.4) | 7082 (37.5) | 7134 (37.8) | 5157 (38.6) | 10281 (37.4) | 4023 (37.9) | 3976 (37.4) |
| Excessive Drinker | 7614 (24.4) | 6238 (22.7) | 4464 (23.6) | 4673 (24.7) | 2908 (21.7) | 6238 (22.7) | 2414 (22.7) | 2368 (22.3) |
| Missing | 7148 (22.9) | 6795 (24.7) | 4475 (23.7) | 4232 (22.4) | 3299 (24.7) | 6795 (24.7) | 2599 (24.5) | 2771 (26.1) |
| BMI, mean (SD) | 30.3 (6.4) | 28.44 (5.6) | 29.36 (6.0) | 29.48 (5.7) | 30.37 (6.2) | 28.44 (5.6) | 29.58 (5.9) | 29.65 (5.9) |
| BMI Categories, n(%) |  |  |  |  |  |  |  |  |
| Underweight (<18.5) | 173 (0.6) | 343 (1.3) | 151 (0.8) | 105 (0.6) | 67 (0.5) | 343 (1.3) | 65 (0.6) | 62 (0.6) |
| Underweight (<25) | 5597 (17.9) | 6898 (25.1) | 4059 (21.5) | 3645 (19.3) | 2243 (16.8) | 6898 (25.1) | 2074 (19.5) | 2033 (19.1) |
| Overweight (25-30) | 10948 (35.1) | 10536 (38.3) | 7156 (37.9) | 7171 (38.0) | 4718 (35.3) | 10536 (38.3) | 4055 (38.2) | 3894 (36.7) |
| Obese (>30) | 13300 (42.6) | 8470 (30.8) | 6771 (35.8) | 7271 (38.5) | 5854 (43.8) | 8470 (30.8) | 4004 (37.7) | 4185 (39.4) |
| Missing | 1176 (3.8) | 1253 (4.6) | 758 (4.0) | 703 (3.7) | 495 (3.7) | 1253 (4.6) | 425 (4.0) | 449 (4.2) |
|  | ACE-I<br>(n=31,194) | CCB<br>(n=27,500) | ACE-I<br>(n=18,895) | CCB<br>(n=18,895) | ARB<br>(n=13,377) | CCB<br>(n=27,500) | ARB<br>(n=10,623) | CCB<br>(n=10,623) |
| Primary Analysis Cohort |  |  |  |  |  |  |  |  |
| Hypertension duration,<br>mean years (SD) | 9.8 (7.4) | 8.6 (7.5) | 9.4 (6.9) | 9.4 (8.0) | 11.4 (7.5) | 8.6 (7.5) | 10.5 (7.0) | 11.3 (8.3) |
| Age at hypertension diagnosis,<br>mean (SD) | 51.0 (10.9) | 58.8 (11.2) | 55.3 (10.4) | 54.0 (9.1) | 52.4 (10.9) | 58.8 (11.2) | 54.5 (10.4) | 53.2 (11.0) |
| Systolic BP, mean(SD) | 135.9 (13.8) | 137.3 (13.6) | 136.7 (14.0) | 136.4 (13.1) | 136.3 (13.6) | 137.3 (13.6) | 136.8 (13.6) | 136.6 (13.4) |
| Diastolic BP, mean(SD) | 81.7 (9.4) | 79.76 (9.3) | 80.6 (9.3) | 81.09 (9.0) | 80.8 (9.1) | 79.8 (9.3) | 80.5 (9.2) | 80.5 (9.3) |
| Missing, n (%) | 80 (0.3) | 63 (0.2) | 0 (0%) | 0 (0%) | 22 (0.2) | 63 (0.2) | 0 (0%) | 0 (0%) |
| Cholesterol, mean (SD) | 5.13 (1.03) | 5.13 (1.04) | 5.12 (1.04) | 5.13 (1.04) | 5.13 (1.01) | 5.13 (1.04) | 5.14 (1.02) | 5.13 (1.03) |
| Cholesterol Categories, n (%) |  |  |  |  |  |  |  |  |
| <5.2 mmol/L | 17051 (54.7) | 14915 (54.2) | 10430 (55.2) | 10394 (55.0) | 7415 (55.4) | 14915 (54.2) | 5820 (54.8) | 5894 (55.5) |
| 5.2-6.2 mmol/L | 8852 (28.4) | 7712 (28.0) | 5262 (27.9) | 5323 (28.2) | 3836 (28.7) | 7712 (28.0) | 3026 (28.5) | 2929 (27.6) |
| >=6.2 mmol/L | 4153 (13.3) | 3747 (13.6) | 2543 (13.5) | 2580 (13.7) | 1699 (12.7) | 3747 (13.6) | 1419 (13.4) | 1404 (13.2) |
| Missing | 1138 (3.7) | 1126 (4.1) | 660 (3.5) | 598 (3.2) | 427 (3.2) | 1126 (4.1) | 358 (3.4) | 396 (3.7) |
| HDL, mean(SD) | 1.44 (0.43) | 1.54 (0.46) | 1.49 (0.44) | 1.48 (0.44) | 1.48 (0.43) | 1.54 (0.46) | 1.50 (0.44) | 1.50 (0.45) |
| HDL categories, n(%) |  |  |  |  |  |  |  |  |
| <1.55 mmol/L | 19720 (63.2) | 14873 (54.1) | 10949 (58.0) | 11464 (60.7) | 8160 (61.0) | 14873 (54.1) | 6214 (58.5) | 6251 (58.8) |
| >=1.55 mmol/L | 9851 (31.6) | 11114 (40.4) | 7007 (37.1) | 6566 (34.8) | 4621 (34.5) | 11114 (40.4) | 3915 (36.9) | 3818 (35.9) |
| Missing | 1623 (5.20) | 1513 (5.5) | 939 (5.0) | 865 (4.6) | 596 (4.5) | 1513 (5.5) | 494 (4.7) | 554 (5.2) |
| eGFR, mean(SD) | 86.1 (14.7) | 82.8 (13.8) | 83.33 (14.3) | 85.08 (14.0) | 82.9 (14.6) | 82.8 (13.8) | 82.36 (14.2) | 83.89 (14.7) |
| eGFR category, n(%) |  |  |  |  |  |  |  |  |
| >60 (Stage 2 and above) | 29735 (95.3) | 25680 (93.4) | 17806 (94.2) | 17924 (94.9) | 12538 (93.7) | 25680 (93.4) | 9981 (94.0) | 9927 (93.5) |
| 30-59(Stage 3) | 1127 (3.6) | 1270 (4.6) | 847 (4.5) | 792 (4.2) | 713 (5.3) | 1270 (4.6) | 530 (5.0) | 578 (5.4) |
| Missing | 332 (1.1) | 550 (2.0) | 242 (1.3) | 179 (1.0) | 126 (0.9) | 550 (2.0) | 112 (1.1) | 118 (1.1) |
| Baseline conditions, n (%) |  |  |  |  |  |  |  |  |
| Atrial fibrillation | 726 (2.3) | 743 (2.7) | 503 (2.7) | 502 (2.7) | 384 (2.9) | 743 (2.7) | 318 (3.0) | 332 (3.1) |
| Rheumatoid arthritis | 460 (1.5) | 497 (1.8) | 329 (1.7) | 297 (1.6) | 199 (1.5) | 497 (1.8) | 161 (1.5) | 166 (1.6) |
| Cancer | 2525 (8.1) | 3320 (12.1) | 1921 (10.2) | 1711 (9.1) | 1377 (10.3) | 3320 (12.1) | 1151 (10.8) | 1139 (10.7) |
| Respiratory Disease | 1325 (4.3) | 1695 (6.2) | 1010 (5.4) | 933 (4.9) | 664 (5.0) | 1695 (6.2) | 586 (5.5) | 572 (5.4) |
|  | ACE-I<br>(n=31,194) | CCB<br>(n=27,500) | ACE-I<br>(n=18,895) | CCB<br>(n=18,895) | ARB<br>(n=13,377) | CCB<br>(n=27,500) | ARB<br>(n=10,623) | CCB<br>(n=10,623) |
| Primary Analysis Cohort |  |  |  |  |  |  |  |  |
| Immunosuppressed* | 571 (1.8) | 616 (2.2) | 392 (2.1) | 358 (1.9) | 239 (1.8) | 616 (2.2) | 205 (1.9) | 198 (1.9) |
| Rare metabolic disorder | 29 (0.1) | 38 (0.1) | 23 (0.1) | 18 (0.1) | 18 (0.1) | 38 (0.1) | 15 (0.1) | 13 (0.1) |
| Other medications at baseline, n (%) |  |  |  |  |  |  |  |  |
| Previous prescription of ACE-I | 31194 (100) | 6030 (21.9) | 18895 (100) | 4462 (23.6) | 8421 (63.0) | 6030 (21.9) | 5686 (53.5) | 5638 (53.1) |
| Previous prescription of ARB | 1036 (3.3) | 1907 (6.9) | 937 (5.0) | 762 (4.0) | 13377 (100) | 1907 (6.9) | 10623 (100) | 1492 (14.0) |
| Previous prescription of CCB | 8242 (26.4) | 27500 (100) | 5554 (29.4) | 18895 (100) | 5393 (40.3) | 27500 (100) | 4001 (37.7) | 10623 (100) |
| Thiazide diuretics | 5775 (18.5) | 3919 (14.3) | 5775 (18.5) | 3919 (14.3) | 3457 (25.8) | 3919 (14.3) | 2074 (19.5) | 2468 (23.2) |
| Loop diuretics | 768 (2.5) | 461 (1.7) | 768 (2.5) | 461 (1.7) | 417 (3.1) | 461 (1.7) | 331 (3.1) | 219 (2.1) |
| Potassium diuretics | 148 (0.5) | 101 (0.4) | 148 (0.5) | 101 (0.4) | 85 (0.6) | 101 (0.4) | 59 (0.6) | 69 (0.7) |
| Alpha blockers | 1017 (3.3) | 659 (2.4) | 1017 (3.3) | 659 (2.4) | 661 (4.9) | 659 (2.4) | 430 (4.1) | 481 (4.5) |
| Beta blockers | 3525 (11.3) | 3030 (11.0) | 3525 (11.3) | 3030 (11.0) | 1630 (12.2) | 3030 (11.0) | 1302 (12.3) | 1423 (13.4) |
| Other antihypertensive | 122 (0.4) | 84 (0.3) | 122 (0.4) | 84 (0.3) | 71 (0.5) | 84 (0.3) | 48 (0.5) | 48 (0.5) |
| Anticoagulants | 884 (2.8) | 889 (3.2) | 884 (2.8) | 889 (3.2) | 446 (3.3) | 889 (3.2) | 370 (3.5) | 385 (3.6) |
| Statins | 9399 (30.1) | 9133 (33.2) | 9399 (30.1) | 9133 (33.2) | 3977 (29.7) | 9133 (33.2) | 3293 (31.0) | 3230 (30.4) |

\*(Treatment with immunosuppressive therapies/antibody treatment/solid organ transplant/ chemo/radiotherapies)
ACE-I=angiotensin converting enzyme inhibitor, ARB=angiotensin II receptor blocker, CCB=calcium channel blocker

Duration of hypertension was slightly longer for users of ACE inhibitors than CCBs (9.8 years vs 8.6 years, respectively). Systolic and diastolic BP, cholesterol, and renal function were similar across both groups. The prevalence of comorbidities was also similar between groups, except for cancers and respiratory disease, which were slightly more common in the CCB cohort.

21.9% of the CCB cohort had previously used an ACE inhibitor and 26.4% of the ACE inhibitor cohort had previously used a CCB. Prescription of other antihypertensives was similar between groups except for thiazide diuretics, which was slightly more common in the ACE inhibitor cohort (18.5% in users of ACE inhibitors vs 14.3% in users of CCBs). Prescriptions of statins were also similar between both cohorts.

Following propensity score matching, users of ACE inhibitors and CCBs were similar in age (64.7 years vs 63.4 years, respectively). Other characteristics were well balanced, including demographic, behavioural and metabolic risk factors, comorbidities and prescriptions (Table1).

#### Angiotensin II receptor blockers versus calcium channel blockers

Before matching, the mean age of users of ARBs was younger than users of CCBs (63.8 years vs 67.4, respectively [Table 1]). However, the proportion of males was slightly lower in users of ARBs than CCBs (40.2% vs 45.6%, respectively). A smaller proportion of users of ARBs were current smokers compared to users of CCBs (7.9% vs 12.7%, respectively). A greater proportion of users of ARBs were overweight or obese than users of CCBs (79.1% vs 69.1%).

The ARB cohort had a longer mean duration of hypertension than the CCB cohort (11.4 years vs 8.6, respectively) and a younger mean age at hypertension diagnosis (52.4 years vs 58.8). BP, cholesterol and renal function were similar between groups, as was the prevalence of comorbidities.

63.0% of users of ARBs had previously used an ACE inhibitor, compared to 21.9% of users of CCBs. 40.3% of the ARB cohort had previously used a CCB while 6.9% of the CCB cohort had previously used an ARB. Users of ARBs were more likely than users of CCBs to have been prescribed thiazide diuretics as well as other antihypertensive drugs. However, the proportion with a prescription for statins was slightly greater in the CCB cohort.

Following propensity score matching, characteristics were well balanced between both groups, including demographic characteristics, behavioural risk factors, metabolic profile, comorbidities, and prescriptions (Table 1).

### Outcomes

#### ACE inhibitors versus calcium channel blockers

Before matching, 148 individuals (0.47%) in the ACE inhibitor cohort developed suspected or confirmed COVID-19 during 14,733 person-years of follow-up, representing a crude incidence rate of 10.1 per 1000 person-years (Table 2). 126 individuals (0.46%) in the CCB cohort developed suspected or confirmed COVID-19 during 12,985 person-years, representing a crude incidence rate of 9.70 per 1000 person-years in the CCB cohort. The unadjusted hazard ratio for suspected/confirmed COVID-19 comparing the ACE inhibitor cohort to the CCB cohort (as the reference) was 1.04 (95% CI 0.82 to 1.31), which fell slightly to 1.01 (95% 0.78 to 1.30) after adjusting for measured confounders.

**Table 2:** Main outcomes for the primary, secondary, sensitivity and negative outcome analyses

|  | Primary Analysis |  | Sensitivity Analysis |  | Primary Analysis |  | Sensitivity Analysis |  |
| --- | --- | --- | --- | --- | --- | --- | --- | --- |
|  | ACE-I | CCB | ACE-I<br>+/- CCB | CCB | ARB | CCB | ARB<br>+/- CCB | CCB |
| **Suspected or confirmed COVID-19** |  |  |  |  |  |  |  |  |
| ***Propensity score matched analysis*** |  |  |  |  |  |  |  |  |
| Outcome events, n (%) | 83 (0.44) | 85 (0.45) | 108 (0.45) | 108 (0.45) | 79 (0.74) | 58 (0.55) | 88 (0.66) | 75 (0.57) |
| Person-years | 8,923 | 8,932 | 11,211 | 11,219 | 5,010 | 5,016 | 6,258 | 6,267 |
| Crude incidence rate/1000 person-years | 9.30 | 9.52 | 9.63 | 9.63 | 15.77 | 11.56 | 14.06 | 11.97 |
| Unadjusted incidence rate ratio (95% CI) | 0.98 (0.72-1.32) p=0.885 |  | 1.00 (0.77-1.31) p=0.994 |  | 1.36 (0.97-1.91) p=0.072 |  | 1.18 (0.86-1.60) p=0.304 |  |
| Adjusted hazard ratio (95% CI) | 0.92 (0.68-1.26) p=0.609 |  | 0.95 (0.72-1.24) p=0.703 |  | 1.38 (0.98-1.95) p=0.062 |  | 1.18 (0.86-1.61) p=0.303 |  |
| **All-cause mortality** |  |  |  |  |  |  |  |  |
| ***Propensity score matched Analysis*** |  |  |  |  |  |  |  |  |
| Outcome events, n (%) | 102 (0.54) | 66 (0.35) | 131 (0.55) | 109 (0.46) | 40 (0.38) | 50 (0.47) | 55 (0.41) | 70 (0.53) |
| Person-years | 8,941 | 8,950 | 11,234 | 11,242 | 5,028 | 5,028 | 6,278 | 6,282 |
| Crude incidence rate/1000 person years | 11.41 | 7.37 | 11.66 | 9.70 | 7.96 | 9.94 | 8.76 | 11.14 |
| Unadjusted incidence rate ratio (95% CI) | 1.55 (1.14-2.11) p=0.006 |  | 1.20 (0.93-1.55) p=0.155 |  | 0.80 (0.53-1.21) p=0.293 |  | 0.79 (0.55-1.12) p=0.182 |  |
| Adjusted hazard ratio (95% CI) | 1.25 (0.90-1.73) p=0.182 |  | 1.00 (0.77-1.30) p=0.977 |  | 0.85 (0.56-1.30) p=0.458 |  | 0.76 (0.53-1.09) p=0.138 |  |
| **Negative Control Outcome**<br>**(Trauma/Accidents/Injury)** |  |  |  |  |  |  |  |  |
| ***PS Matched Analysis*** |  |  |  |  |  |  |  |  |
| Outcome events, n (%) | 185 (0.98) | 187 (0.99) | 246 (1.04) | 246 (1.04) | 105 (0.99) | 108 (1.02) | 137 (1.03) | 146 (1.10) |
| Person-years | 8,898 | 8,904 | 11,176 | 11,182 | 5,004 | 5,002 | 6,245 | 6,246 |
| Crude incidence rate/1000 person years | 20.79 | 21.00 | 22.01 | 22.00 | 20.98 | 21.59 | 21.94 | 23.37 |
| Unadjusted incidence rate ratio (95% CI) | 0.99 (0.81-1.21) p=0.923 |  | 1.00 (0.84-1.19) p=0.996 |  | 0.97 (0.74-1.27) p=0.835 |  | 0.94 (0.74-1.18) p=0.594 |  |
| Adjusted hazard ratio (95% CI) | 0.95 (0.77-1.17) p=0.640 |  | 0.96 (0.80-1.14) p=0.620 |  | 0.97 (0.74-1.27) p=0.801 |  | 0.91 (0.72-1.15) p=0.439 |  |

Following propensity score matching, 83 individuals (0.44%) in the ACE inhibitor cohort had suspected or diagnosed COVID-19 during 8923 person-years of follow-up, representing a crude incidence rate of 9.30 per 1000 person-years. 85 individuals (0.45%) in the CCB cohort developed suspected or confirmed COVID-19 during 8932 person-years of follow-up representing a crude incidence rate of 9.5 per 1000 person-years. The unadjusted hazard ratio for suspected/confirmed COVID-19 comparing the ACE inhibitor cohort to the CCB cohort was 0.98 (95% CI 0.72 to 1.32). Upon adjustment for measured confounders, the hazard ratio was 0.92 (95% 0.68 to 1.26; Figure 2).

**Figure 2:**
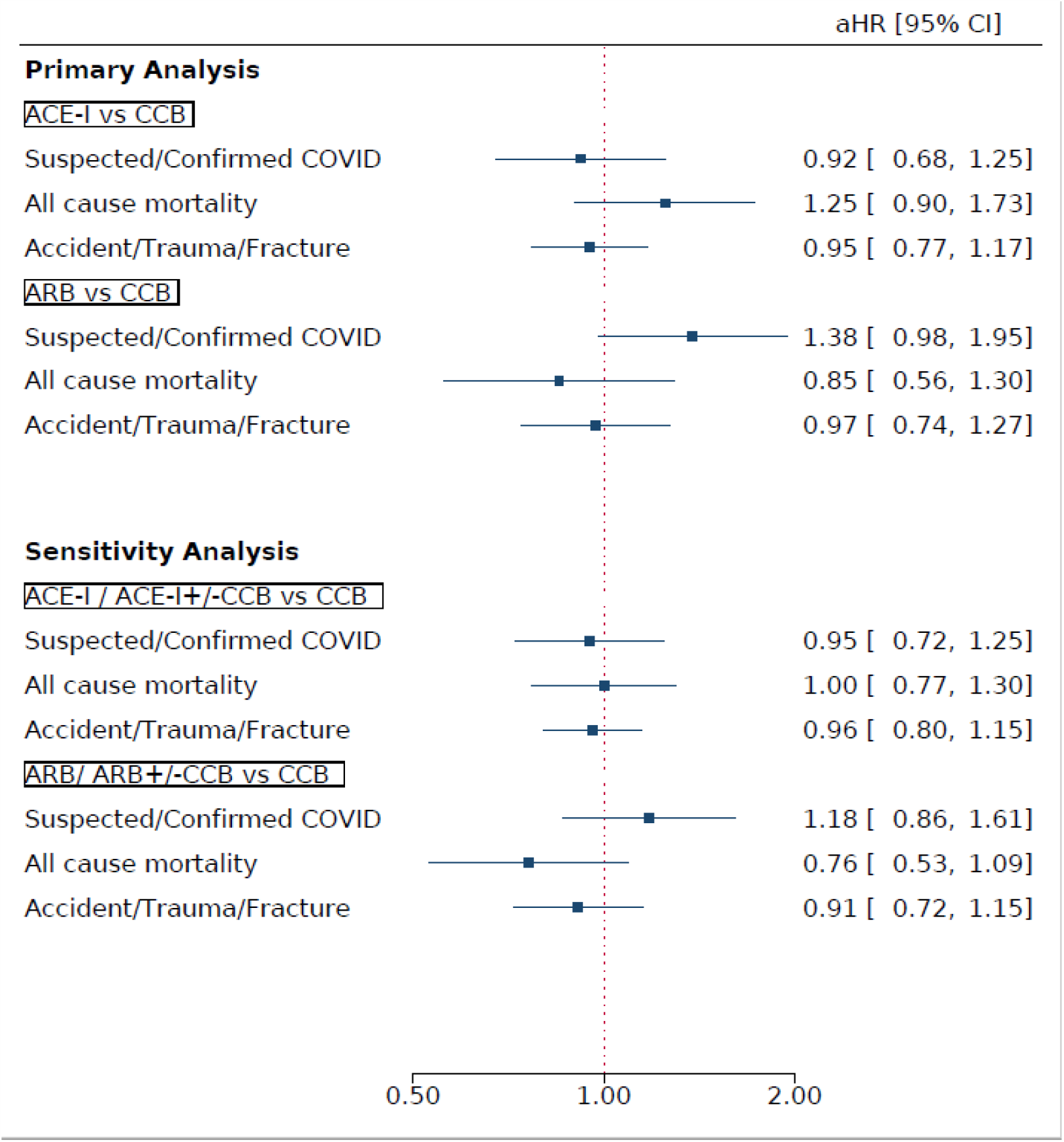
Forest plot of adjusted hazard ratios for suspected or confirmed COVID-19, all-cause mortality and accidents, trauma and fractures (negative control) (ACE-I=angiotensin converting enzyme inhibitor, ARB=angiotensin II receptor blocker, CCB=calcium channel blocker)

Similar results were found in the sensitivity analyses for the primary outcome. When comparing hypertensive users of ACE inhibitors with or without CCBs to those using CCBs alone, the adjusted hazard ratio was 0.95 (95% CI 0.72 to 1.25) following propensity score matching. When including individuals with diabetes mellitus, ischemic heart disease, stroke, transient ischaemic attack and peripheral vascular disease, the adjusted hazard ratio for COVID-19 after propensity score matching was 0.91 (95% CI 0.74 to 1.12). When including individuals with any comorbidities, the adjusted hazard ratio after propensity score matching was similarly 0.98 (95% CI 0.81 to 1.18; Figure 3).

**Figure 3:**
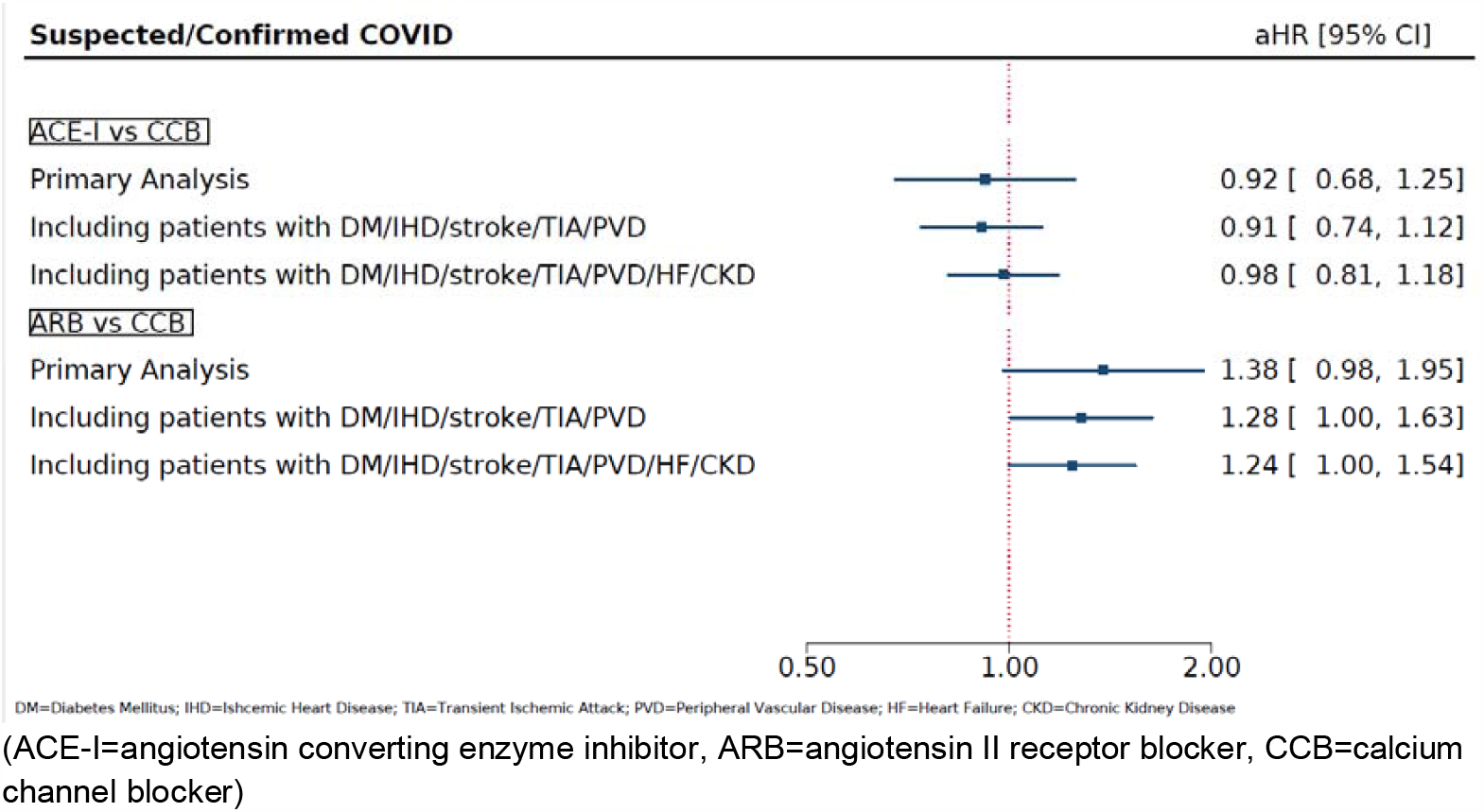
Forest plot of hazard ratios for suspected or confirmed COVID-19 when including comorbidities that were excluded in the primary analysis. (ACE-I=angiotensin converting enzyme inhibitor, ARB=angiotensin II receptor blocker, CCB=calcium channel blocker)

The propensity score-matched analysis for all-cause mortality produced a statistically non-significant adjusted hazard ratio of 1.25 (95% CI 0.90 to 1.73). The negative control analysis similarly found no statistically significant association between prescription of ACE inhibitors and accidents, trauma or fractures compared to prescription of CCBs (adjusted HR 0.95 (95% CI 0.77 to 1.17).

#### Angiotensin II receptor blockers versus calcium channel blockers

Before matching, 99 individuals (0.74%) in the ARB cohort developed suspected or confirmed COVID-19 over a follow-up of 6308 person-years, representing a crude incidence rate of 15.7 per 1000 person-years. 126 (0.46%) in the CCB cohort developed suspected or confirmed COVID-19 over a follow-up of 12,985 person-years, representing a crude incidence rate of 9.7 per 1000 person-years. The unadjusted hazard ratio for suspected or confirmed COVID-19 when comparing the ARB cohort to the CCB cohort (as the reference) was 1.62 (95% CI 1.24 to 2.10). After adjustment for measured confounders, the hazard ratio was slightly attenuated to 1.51 (95% CI 1.12 to 2.03), although remained statistically significant.

After propensity score matching, 79 individuals (0.74%) in the ARB cohort developed suspected or confirmed COVID-19 over 5010 person-years of follow-up, representing a crude incidence rate of 15.8 per 1000 person-years. In the CCB cohort, 58 individuals (0.55%) developed suspected or confirmed COVID-19 over 5016 years of follow-up, representing a crude incidence rate of 11.6 per 1000 person-years. The unadjusted hazard ratio when comparing the ARB cohort to the CCB cohort was 1.36 (95% CI 0.97 to 1.91). After adjustment for measured confounders, the hazard ratio increased slightly to 1.38 (95% CI 0.98 to 1.95; Figure 2).

These findings were attenuated in sensitivity analyses when comparing users of ARBs with or without concurrent use of CCBs to propensity score-matched individuals using CCBs alone (adjusted HR 1.18, 95% CI 0.86 to 1.61). When only excluding individuals with either heart failure or CKD, the adjusted hazard ratio was slightly attenuated compared to the primary analysis but statistically significant after propensity score matching (adjusted HR 1.28, 95% CI 1.00 to 1.63). This was similarly found when including propensity score-matched individuals with any comorbidity (adjusted HR 1.24, 95% CI 1.00 to 1.54; Figure 3).

There was no statistically significant difference in all-cause mortality between users of ARBs compared to users of CCBs after propensity score matching (adjusted HR 0.85, 95% CI 0.56 to 1.30). Similarly, there was no association between use of ARBs and the negative control outcome of accidents, trauma and fractures when compared to propensity score matched users of CCBs (adjusted HR 0.97, 95% CI 0.74 to 1.27).

## Discussion

### Main findings

We found no difference in the risk of developing suspected or confirmed COVID-19 or all-cause mortality among individuals with hypertension treated with ACE inhibitors compared to those treated with CCBs, after matching and adjusting for a wide range of risk factors known to be associated with COVID-19, as well as indications for ACE inhibitor prescription. However, we found a statistically non-significant 38% relative increase in risk of the development of suspected or confirmed COVID-19 among those prescribed ARBs compared to those prescribed CCBs, but no difference in all-cause mortality.

### Relationship to other studies

There has been ongoing debate over whether RAS inhibitors are protective or harmful in the context of COVID-19. Concerns were raised early on in the pandemic speculating that this class of drugs could increase susceptibility to COVID-19 by upregulating ACE2 receptors, and thus promoting entry of SARS-CoV-2 virus into host cells.(19) Two *in vivo* studies in rats showed that ACE inhibitors increased ACE2 activity in the plasma and renal cortex.(20)(21) However, these studies involved far higher doses of ACE inhibitors than would typically be used in humans. In fact, a review of 11 human studies overwhelmingly showed that RAS inhibitors do not increase plasma or urine ACE2 expression, (Sriram and Insel 2020) although it remains unknown whether there is any effect on membrane-bound ACE2 activity. Indeed, there are no studies to-date on the effects of RAS inhibitors specifically on lung ACE2 expression.(22)(10)

Once within the cell, coronaviruses themselves downregulate ACE2 expression in host cells, which is understood to reduce the pulmonary activity of the anti-inflammatory ACE2/angiotensin 1-7/mas receptor system.(19)(7) This results in angiotensin II proliferation and consequent lung inflammation. In one small study of hospitalised patients with COVID-19, angiotensin II levels were markedly elevated and linearly associated with viral load and severity of lung injury.(23) Both ACE inhibitors and ARBs have been shown to attenuate the inflammatory response in mouse models, potentially through the inhibition of interleukin 6 (IL-6).(24)

It is possible that RAS inhibitors confer a protective effect through their anti-inflammatory actions, although further evidence of such a mechanism is needed. One study found that an ARB attenuated acute pulmonary oedema and lung damage in mice infected with SARS-CoV, which is a close relative of SARS-CoV-2.(7) Two small case series in humans both found that SARS-CoV-2 infected patients on RAS inhibitors had significantly lower inflammatory markers, and increased CD3 and CD8 T cell proliferation than patients on alternative antihypertensive medications.(7)(25)(26)

More recently, meta-analyses of observational studies have also shown that RAS inhibitors are not associated with severe outcomes and death in hospitalised patients with COVID-19. Grover et al. identified 16 studies comparing clinical outcomes and mortality in inpatients with COVID-19 who had been prescribed ACE inhibitors and/or ARBs. In a pooled analysis, RAS inhibitor use was non-significantly associated with lower odds of developing severe disease (OR 0.81, 95% CI 0.41 to 1.58) and mortality (OR 0.86, 95% CI 0.53 to 1.41).(27) Similarly, in a pooled analysis of 11 studies, Pranata et al. found a non-significantly lower adjusted odds of mortality in those on RAS inhibitors (OR 0.83, 95% CI 0.54 to 1.27) but no difference for disease severity (OR 1.03, 95% CI 0.73 to 1.45).(28) In contrast to our findings, a subgroup analysis found that ARBs were associated with reduced mortality (OR 0.51, 95% CI 0.29 to 0.90) but the association was not statistically significant for ACE inhibitors (OR 0.68, 95% CI 0.39 to 1.17).

A meta-analysis of five observational studies by Adbulhak et al. also showed a reduced risk from critical or fatal outcomes among patients with COVID-19 who took RAS inhibitors, with a pooled odds ratio of 0.32 (95% CI 0.22 to 0.46).(29) Ghosal et al. similarly showed a reduction in the odds of severe disease and death (OR 0.56 for severe illness, 95% CI 0.34 to 1.89, and OR 0.38 for death, 95% CI 0.19 to 0.74).(30) A systematic review of RAS inhibitors and COVID-19 by Zhang et al. also showed that their use was not associated with increased likelihood of testing positive for SARS-CoV-2 or with severity of disease once infected.(31)

The studies included in these meta-analyses were largely based on hospital cohorts in contrast to our study which included patients registered in primary care. Prescriptions of and compliance with pre-hospital medications may be much better recorded in primary care cohorts. Hospital-based cohorts typically include the most severely ill patients and do not include those with asymptomatic or mild-to-moderate disease. Furthermore, many of the included studies did not adjust effect estimates for potential confounding factors or assess the effects of sub-classes of RAS inhibitors (ACEIs and ARBs) separately, which our study suggests may not be uniform.

Mancia et al. conducted a large population-based study in Lombardy of those diagnosed with COVID-19 matched to population controls on age, sex and geography.(15) After multivariable adjustment, neither ACE inhibitors or ARBs showed an association with the risk of developing COVID-19 (OR 0.95 [95% CI 0.86 to 1.05] and 0.96 [95% CI 0.87 to 1.07], respectively. Another large retrospective cohort and nested case-control study of all Danish people assessed in hospital with COVID-19 in a three month period found no association between ACE inhibitor/ARB use susceptibility to COVID-19 or mortality when compared with other antihypertensives.(16)

A large multinational cohort study, which used propensity score matching and negative controls found, as we did, that prescription of ACE inhibitors or ARBs was not associated with the risk of diagnosis of COVID-19 in comparison to use of calcium channel blockers or thiazide diuretics.(32) When directly comparing invididuals prescribed ACE inhibitors with those prescribed ARBs, there was a higher risk of COVID-19 diagnosis in the latter. However, there were no significant differences in COVID-19 related hospitalisation between all antihypertensive drug classes.

Most recently, a prospective cohort study using data from general practices in England found that use of ACE inhibitors or ARBs was associated with a significantly reduced risk of COVID-19 but not associated ICU admission.(33) This study also found that ethnicity modified the association between use of RAS inhibitors and COVID-19, with those from Black ethnic groups being at increased risk, a trend we were not able to explore in our study.

The above studies included patients with a range of comorbidities that were potential indications for RAS inhibitors and were therefore potentially prone to prescription by indication bias. We limited our inclusion criteria to patients with hypertension and excluded those with other conditions that were potential indications for RAS inhibitors in our main analysis to limit the effect of these biases. However, our findings remained largely in line with these prior studies.

### Strengths and limitations

The primary outcome of suspected or confirmed COVID-19 may not have been well recorded in primary care records. Relatively little testing for COVID-19 occurred early in the pandemic and data flows from COVID-19 testing centres and hospitals to primary care has generally been suboptimal. However, we expect this effect to have been equally distributed across all our included drug exposure cohorts and should therefore not have biased our effect estimates. We also did not have access to data on hospitalisations or cause-specific mortality. Because of the low numbers of deaths in each drug exposure cohort, we did not have sufficient statistical power to assess the association between drug exposures and COVID-19 mortality. We also had insufficient data on ethnicity and socioeconomic status to include this in our analyses, both of which are known to be associated with COVID-19.

The strengths of the study include the study design, which attempted to control for confounding by indication bias and adjust for a large number of known risk factors for COVID-19. We also performed multiple sensitivity analyses to check the robustness of our findings in comparison with other published studies.

### Implications for practice, policy and research

Despite initial concerns about the safety of ACE inhibitors in the context of SARS-CoV-2 pathophysiology, they appear to have no effect on susceptibility to COVID-19 compared to the use of CCBs. Our findings should provide further reassurance, in addition to previously published studies on this topic, that prescription of ACE inhibitors does not increase vulnerability to being infected with SARS-CoV-2.

However, our findings suggest that the effects of RAS inhibitors are not uniform across drug classes. ARBs by contrast were associated with a statistically non-significant increased risk of presentation with COVID-19, but not mortality, in comparison to the use of CCBs. The reasons for this are unclear but one potential reason may be due to differential health-seeking behaviour between users of ACE inhibitors and ARBs. One common side effect of ACE inhibitors is cough due to the reduced breakdown of bradykinin and substance P, which are degraded by ACE, and a rise in prostaglandins due to increased concentrations of bradykinin.(34) Patients who experience cough secondary to ACE inhibitors are frequently switched to ARBs, and indeed a large proportion of patients in our ARB cohort had been previously prescribed an ACE inhibitor. Those prescribed ARBs could therefore be more prone to coughing, which could, in turn, have made them more likely to present to healthcare services with symptoms of COVID-19. Reassuringly, we did not see any association between use of ARBs and all-cause mortality during the peak of the pandemic, supporting the hypothesis that the association between their use and COVID-19 is likely related to differences in health seeking behaviour rather than a true increase in susceptibility to the infection.

An alternative hypothesis is that the differences in COVID-19 risk observed between ACE inhibitors and ARBs is due to residual confounding. We did not for example have data on ethnicity or socioeconomic status and it is possible that patients from different ethnic groups or social classes could be prescribed antihypertensives differentially and have different patterns of health-seeking behaviour.

Nevertheless, there remains the possibility that the higher risk of COVID-19 observed among users of ARBs is causal and that this class of drugs increases susceptibility to SARS-CoV-2 but not all-cause mortality. Further research is needed to test the biological plausibility and causality of this apparent effect. Further research is also needed to assess whether the use of ACEIs and ARBs is associated with COVID-19 hospitalisation and death and whether any such associations differ between drug classes.

## Conclusions

Prescription of ACE inhibitors was not associated with the risk of suspected or confirmed COVID-19 in primary care. By contrast, prescription of angiotensin II receptor blockers was associated with a statistically non-significant increase in risk. However, neither drug class was associated with all-cause mortality during the peak of the pandemic. These findings need to be confirmed in other observational studies, potential pathways modelled through causal inference studies, and the basic mechanistic science of this potential association to be understood before recommendations can be made on the clinical implications for RAS inhibitor use during the ongoing COVID-19 pandemic.

## Data Availability

The study protocol can be accessed at the European Union electronic Register of Post-Authorisation Studies (EU PAS) register.

## Notes

### Competing Interest Statement

The authors have declared no competing interest.

### Clinical Protocols

http://www.encepp.eu/encepp/viewResource.htm?id=35329

### Funding Statement

This study was not externally funded. ES reports receiving funding from HDR-UK (PIONEER Hub), Wellcome, MRC, British Lung Foundation and NIHR. DP reports receiving funding from NIHR and MRC.

